# Injury epidemiology in HYROX athletes: an international cross-sectional survey

**DOI:** 10.64898/2026.08.09.26359590

**Authors:** Conrad Ketzer, Luca Kirstein, Magdalena Bonleitner, Philipp Beyerle, Philipp Zehnder, Markus Schwarz, Peter Biberthaler, Michael Zyskowski

## Abstract

**Objective:** HYROX is a rapidly growing hybrid fitness competition combining running with functional exercise stations. Our objective was to describe the 12-month prevalence, characteristics and severity of self-reported HYROX-related injuries.

**Methods:** We conducted an international cross-sectional online survey of 418 HYROX athletes. The primary outcome was the self-reported 12-month period prevalence of at least one HYROX-related injury; secondary outcomes included an exposure-adjusted lower-bound rate per 1000 hours of total training exposure and the profile and severity of the most significant injury. Associated factors were examined by multivariable logistic regression.

**Results:** Overall, 208 of 418 participants (49.8%, 95% CI 45.0 to 54.5) reported at least one HYROX-related injury. The exposure-adjusted lower-bound rate was 1.65 reported injuries per 1000 hours of total training exposure. Injuries mainly affected the lower extremity, most commonly the knee (20.8%); tendon-related complaints were the leading type (41.6%) and most were of gradual onset. Among participants with severity data, 20.3% reported more than 28 days of training interruption or no return to their previous performance level. Higher HYROX-specific training frequency was the only factor independently associated with injury reporting (adjusted OR 1.61, 95% CI 1.20 to 2.16; p = 0.001).

**Conclusion:** Approximately half of respondents reported at least one HYROX-related injury during the preceding 12 months, predominantly involving gradual-onset lower-extremity complaints. Higher HYROX-specific training frequency was associated with injury reporting, although the cross-sectional design precludes causal interpretation. Prospective, exposure-based surveillance is needed to quantify HYROX-specific injury incidence and burden and examine whether training frequency, load distribution and recovery contribute to injury risk.

**What is already known on this topic:** HYROX has expanded rapidly as a hybrid-fitness competition format, yet sport- specific injury evidence is limited to a single small cross-sectional study and exposure-adjusted estimates are lacking.

**What this study adds:** Among 418 athletes, self-reported HYROX-related injuries over the preceding 12 months were common (period prevalence 49.8%) and predominantly gradual-onset, lower-extremity and tendon-related. Higher HYROX-specific training frequency was the only measured factor independently associated with injury reporting.

**How this study might affect research, practice or policy:** These hypothesis-generating benchmarks make the case for prospective, exposure- based HYROX injury surveillance; they do not support specific training prescriptions, but identify training frequency, load distribution and recovery as priorities for future study. HYROX’s fixed, reproducible race structure offers a practical real-world model for such research.

## Introduction

Since its launch in Germany in 2017, HYROX has rapidly evolved from a niche event into an international hybrid fitness competition, with more than 1.5 million reported participants across 30 countries by 2026.(1) A race consists of eight 1-km running segments, each followed by one of eight standardised functional stations (SkiErg, sled push, sled pull, burpee broad jumps, rowing, farmer’s carry, sandbag lunges and wall balls).(2) HYROX can therefore be considered a running-focused form of high- intensity functional training, although its fixed structure and integral running component distinguish it from CrossFit and related formats.(3, 4) As participation expands, HYROX has become increasingly relevant to sports medicine, particularly in relation to athlete health and injury prevention.(5, 6)

Despite the rapid expansion of HYROX, the scientific literature has not kept pace. Existing studies have primarily focused on physiological responses, performance determinants and athlete characteristics, whereas injury epidemiology has received little attention.(3, 5, 7) Injury-related evidence is limited to a single cross-sectional study of 80 HYROX athletes, which suggested that injuries may be common but provided limited information on severity or context.(5) Robust sport-specific data on the occurrence, profile and context of injuries are therefore lacking, and findings from running or other functional-fitness formats may not be directly transferable.(8)

We therefore conducted a cross-sectional survey of HYROX athletes, providing, to our knowledge, the first international epidemiological description of HYROX-related injuries. The primary aim was to estimate the self-reported 12-month period prevalence; secondary aims were to derive an exposure-adjusted lower-bound estimate of reported injuries per 1000 hours of total training exposure, to characterise the injury profile (body region, type, mode of onset, severity, time loss and context) and to explore athlete- and training-related factors associated with injury reporting. By providing initial reference estimates, the study was designed to inform future prospective surveillance and hypothesis-driven prevention research.(9)

## Methods

### Study design and reporting

This international cross-sectional study used an anonymous online questionnaire to assess self-reported HYROX-related physical complaints or injuries over the preceding 12 months. Reporting followed the STROBE statement and the STROBE Extension for Sport Injury and Illness Surveillance (STROBE-SIIS).(10, 11) The study was approved by the Ethics Committee of the Technical University of Munich (2026- 32-S-CT) and conducted in accordance with the Declaration of Helsinki; all participants provided electronic informed consent.

### Setting, participants and eligibility

Participants were recruited between February and May 2026 via HYROX-affiliated gyms and coaches, social media, athlete networks and printed flyers. Administration and reporting followed CHERRIES guidance for web-based surveys.(12) Because survey links were open and shareable without visitor tracking, view and participation rates could not be determined; survey flow is reported from questionnaire submission onward. Eligible participants were adults (≥18 years) who had competed in HYROX or undertaken HYROX-specific training, defined as combined running and functional- station workouts modelled on race demands; no minimum duration of HYROX participation or training was required. Responses were excluded if consent was withheld or neither competition nor HYROX-specific training was reported. This yielded a self-selected convenience sample. No formal a priori sample-size calculation was performed; recruitment was guided by a pragmatic feasibility target of approximately 500 responses.

### Survey instrument and data collection

Data were collected using a study-specific questionnaire hosted on SoSci Survey (SoSci Survey GmbH, Munich, Germany). The questionnaire was available in German and English; the English version was AI-translated and reviewed by the study team without formal back-translation. It covered demographics, sporting background, HYROX participation and training characteristics, and HYROX-related physical complaints or injuries during the preceding 12 months. Training-related items included total and HYROX-specific weekly training volume, training frequency, self-rated intensity, endurance–strength balance and coaching or supervision. Before launch, the questionnaire was pilot-tested with 15 individuals to assess comprehensibility, usability and completion time; pilot responses were excluded. The full questionnaire and a completed CHERRIES checklist are provided in the online supplementary material.

### Patient and public involvement

HYROX athletes were involved only as anonymous survey respondents and were not involved in setting the research question, study design, conduct, reporting, or dissemination plans.

### Equity, diversity, and inclusion statement

Eligibility was unrestricted by sex, gender or country of residence, and the survey was available in German and English. Participants represented 30 countries or territories, although the sample was predominantly based in Germany and participation required sufficient German or English proficiency.

### Injury definition, ascertainment and classification

Participants first reported whether they had experienced any HYROX-related complaint or injury in the preceding 12 months (none, one, or more than one). A HYROX-related injury was defined as any physical complaint participants considered related to HYROX training or competition, irrespective of training interruption or medical attention, consistent with the health-problem approach of the International Olympic Committee (IOC).(11) For readability, this outcome is referred to as “injury” hereafter.

Participants described their most significant injury and, where applicable, one further injury, providing detailed data for up to two events per participant. For each event, they reported the context of occurrence and, for injuries during HYROX competition or HYROX-specific training, the HYROX element involved.

Participants further reported body region, injury type, mode of onset, any previous injury or prolonged pain in the same region before the preceding 12 months, severity and healthcare utilisation. Mode of onset was classified as sudden/acute, gradual/overuse, or mixed. Body region categories were adapted from the IOC consensus statement and injury type from its tissue-based categories.(11) Severity was recorded by training interruption: no time loss; 1–3; 4–7; 8–28; >28 days; or no return to the previous performance level.

### Outcomes and exposure

The primary outcome was the self-reported 12-month period prevalence of at least one injury. Secondary outcomes were the profile, severity, context and healthcare utilisation of the most significant injury. Second-injury data were summarised separately and not pooled with the main injury-pattern analyses.

Annual total and HYROX-specific training exposure were estimated from self-reported average weekly training hours (reported as whole hours per week, with the top category ‘>30 h/week’ treated as 31 h/week) multiplied by 52. The exposure-adjusted rate was calculated as the minimum number of reported HYROX-related injuries divided by total (all-activity) training exposure per 1000 hours. Participants reporting more than one injury were conservatively counted as having two; the resulting rate therefore represents a lower-bound estimate.

### Statistical analysis

Statistical analysis and reporting were guided by the CHAMP statement.(13) Continuous variables were summarised as mean (SD) or median (IQR) and categorical variables as counts and percentages. The 12-month period prevalence of at least one injury was reported with a Wilson 95% confidence interval (CI); the exposure-adjusted rate was reported per 1000 hours of total training exposure with an exact Poisson (Garwood) 95% CI.

Factors associated with reporting at least one injury were examined by logistic regression. Univariable models described crude associations, and a multivariable model included a prespecified covariate set: age, self-reported gender, body mass index (BMI), HYROX-specific training frequency and weekly hours, HYROX training experience, self-rated training intensity, coaching or supervision, endurance–strength balance and CrossFit/functional-training background. Univariable and multivariable models used the same complete-case sample (n = 413). HYROX-specific training frequency and experience were analysed as ordinal predictors; corresponding odds ratios represent a one-category increase.

Prevalence ratios were additionally estimated by modified Poisson regression with the same covariates and robust standard errors. Prespecified sensitivity analyses modelled HYROX-specific weekly training volume as tertiles and restricted analyses to fully completed responses and competition-experienced participants. Missing data were handled by complete-case analysis, with the number of observations reported for each analysis. Analyses used Python (pandas, statsmodels, scipy) and IBM SPSS Statistics v29.0 (IBM Corp., Armonk, NY, USA); estimates are reported with 95% CIs, and two-sided p values are interpreted as continuous measures of statistical evidence rather than against a fixed significance threshold.

## Results

### Participants

Between February and May 2026, 560 survey entries were submitted. Three respondents did not provide informed consent and 86 discontinued before eligibility could be assessed. A further 28 reported neither HYROX competition participation nor HYROX-specific training and were therefore ineligible. Of the 443 eligible respondents, 25 discontinued before completing the primary injury outcome item, leaving 418 participants (94.4%) in the main analysis sample (Supplementary Figure 1).

Respondents excluded for non-completion of the primary injury outcome item (n = 25) were younger than analysed participants (30.4 ± 8.9 vs 37.4 ± 9.7 years; p < 0.001) and less likely to report regular HYROX competition participation (8.0% vs 43.1%; p < 0.001), with no clear differences in gender or German versus non-German residence.

### Baseline characteristics

The 418 participants had a mean age of 37.4 ± 9.7 years and 228/417 (54.7%) with available gender data were male. Mean BMI was 24.3 ± 2.8 kg/m². Participants resided in 30 countries or territories, with Germany most frequently represented (274, 65.6%). Sporting backgrounds before HYROX were most commonly strength training (61.5%), endurance sports (51.2%) and CrossFit or functional training (41.6%) (Table 1).

**Table 1:** Baseline characteristics of the study sample (n = 418).

| Characteristic | n = 418 |
| --- | --- |
| **Demographics** |  |
| Age, years — mean ± SD | 37.4 ± 9.7 |
| **Gender — n (%) (n = 417)** |  |
| Male | 228 (54.7) |
| Female | 188 (45.1) |
| Non-binary | 1 (0.2) |
| Height, cm — mean ± SD (n = 417) | 174.6 ± 9.6 |
| Weight, kg — mean ± SD (n = 417) | 75.1 ± 14.0 |
| BMI, kg/m<sup>2</sup> — mean ± SD (n = 414) | 24.3 ± 2.8 |
| **Country or territory of residence — n (%)** |  |
| Germany | 274 (65.6) |
| United States | 31 (7.4) |
| United Kingdom | 22 (5.3) |
| Other (27 countries or territories) | 91 (21.8) |
| **Sporting background before HYROX — n (%)** |  |
| Strength training | 257 (61.5) |
| Endurance sports | 214 (51.2) |
| CrossFit/functional training | 174 (41.6) |
| Team sports | 91 (21.8) |
| Other | 168 (40.2) |

BMI, body mass index; SD, standard deviation. Data are mean ± SD or n (%). Denominators are given where data were missing for individual variables. Percentages are rounded and may not sum to exactly 100%. BMI was calculated from self-reported height and weight; three implausible values (BMI < 15 or > 40 kg/m<sup>2</sup>) were excluded, yielding n = 414. Country or territory of residence was a single-response item with 11 predefined options and a free-text field; free-text entries were harmonised before analysis. "Territories" refers to Hong Kong, reported separately from mainland China. Sporting background reflects a multiple-response item and percentages sum to more than 100%. Other sporting background (n = 168) comprised participants reporting at least one further sport, including outdoor/mountain sports (53), winter sports (43), racket sports (30), combat sports (26), gymnastics/calisthenics (14), athletics (14) and other activities (44); as participants could report several sports, these counts sum to more than 168.

### Training and competition background

Most participants had competition experience: 80.4% had completed at least one HYROX event and 43.1% reported regular competition participation; among competition-experienced participants, 30.7% had competed in the Pro format at least once. HYROX-specific training experience was most commonly 1–2 years (40.7%), with 154 participants (36.8%) reporting less than 12 months. Median total training volume was 8 hours/week (IQR 6–10), including 4 hours/week (IQR 2–6) of HYROX- specific training. HYROX-specific training frequency was most commonly 1–2 days/week (37.8%) or 3–4 days/week (35.2%), with 20.3% training on ≥5 days/week. Median self-rated intensity was 7/10 (IQR 7–8) and the endurance–strength balance score was 41/100 (IQR 28–60) (Supplementary Tables 1–2).

### Injury prevalence and severity

Over the preceding 12 months, 208/418 participants (49.8%; 95% CI 45.0–54.5) reported at least one HYROX-related injury; 123 (29.4%) reported one and 85 (20.3%) more than one. Based on a minimum of 293 injuries over 177,268 hours of total training exposure, the exposure-adjusted lower-bound rate was 1.65 reported injuries per 1000 hours of total training exposure (95% CI 1.47–1.85). Among injured participants with severity data for their most significant injury (n = 197), 179 (90.9%) reported at least one day of interrupted or substantially reduced training and 40 (20.3%) reported >28 days of interruption or no return to their previous performance level. Interruption lasted 1–28 days in 139 participants (70.6%) (Table 2).

**Table 2:** Injury prevalence, exposure-adjusted rate and severity over the preceding 12 months (n = 418).

| Variable | Value |
| --- | --- |
| **Prevalence** |  |
| **Participants reporting ≥1 injury (12-month period prevalence)** | 208/418 (49.8%; 95% CI 45.0–54.5) |
| One injury | 123/418 (29.4%) |
| More than one injury | 85/418 (20.3%) |
| **Exposure-adjusted injury rate** |  |
| Minimum number of reported injuries | 293 |
| Total training exposure, hours | 177,268 |
| Minimum reported injury rate per 1000 training hours | 1.65 (95% CI 1.47–1.85) |
| **Training interruption (most significant injury, n = 197)** |  |
| No interruption (0 days) | 18 (9.1%) |
| 1–3 days | 52 (26.4%) |
| 4–7 days | 41 (20.8%) |
| 8–28 days | 46 (23.4%) |
| > 28 days | 25 (12.7%) |
| No return to previous level yet | 15 (7.6%) |

CI, confidence interval. The 12-month period prevalence is reported with a Wilson 95% CI. The exposure-adjusted lower-bound rate was calculated as the minimum number of reported injuries (participants reporting "more than one" counted as two) divided by total self-reported training exposure, expressed per 1000 hours, with an exact Poisson (Garwood) 95% CI; because the questionnaire did not capture the exact number of injuries among participants reporting more than one, this rate represents an exposure-adjusted lower-bound estimate. Training interruption refers to the most significant reported injury and was available for 197 of 208 injured participants. Categories are mutually exclusive; "no return to previous level yet" denotes injuries from which participants had not yet returned to their pre-injury performance level by the time of survey completion.

### Injury pattern

The knee was the most commonly affected region among the most significant reported injuries (41/197; 20.8%), followed by the spine/back (30/197; 15.2%), lower leg (27/197; 13.7%) and hip/groin (26/197; 13.2%). Tendon-related complaints were the most common injury type (82/197; 41.6%), followed by muscle injuries (39/197; 19.8%) and ligament or joint injuries (26/197; 13.2%). Most injuries had a gradual or mixed onset: 113/197 (57.4%) were attributed to gradual onset or overuse, 44/197 (22.3%) to a mixed onset and 40/197 (20.3%) to an acute onset (Figure 1). A previous injury or prolonged pain in the same region was reported for 29.4% of injuries.

**Figure 1:**
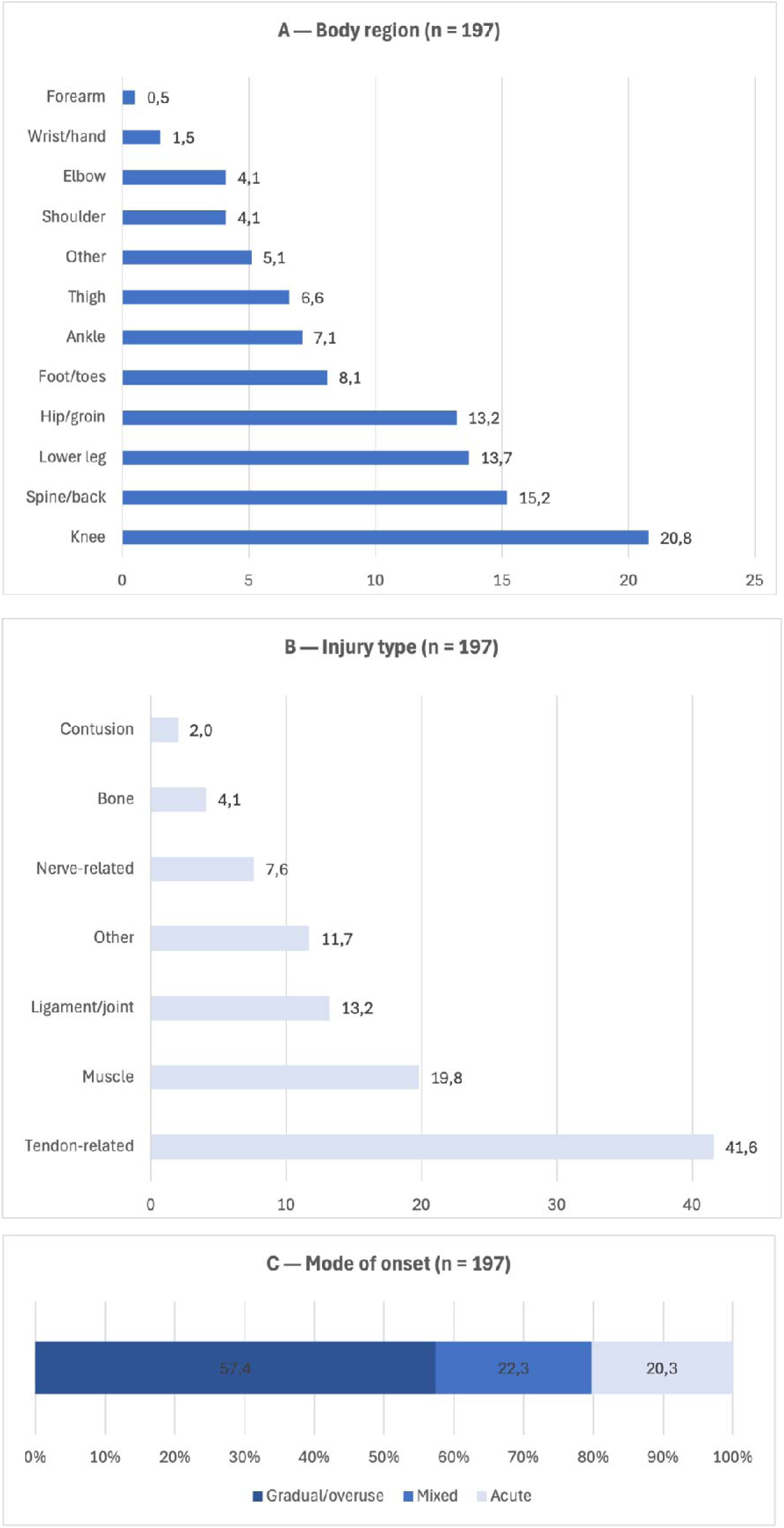
Characteristics of the most significant reported injury (n = 197) (A) Body region, (B) injury type and (C) mode of onset. Panels A and B show the percentage of injuries in each category; panel C shows the distribution of onset as a proportion of all injuries. Percentages are based on the 197 injured participants with available injury-characterisation data. In panel A, “other” comprised regions not captured by the predefined categories, predominantly trunk/thorax complaints; in panel B, “other” comprised heterogeneous free-text entries not corresponding to a single tissue category.

Healthcare use was common: 71.1% reported at least one medical or therapeutic assessment or treatment, most often physiotherapy (53.3%), consultation with an orthopaedic or sports medicine specialist (33.0%) or diagnostic imaging (26.9%) (Supplementary Table 3).

Of the 85 participants reporting more than one injury, 44 provided second-injury details, which showed broadly similar distributions of type, region and onset to the most significant injuries (Supplementary Table 4).

### Context of reported HYROX-related injuries

Of 205 most significant injuries with context data, 37.1% occurred in a directly HYROX-specific setting (28.3% during HYROX-specific training, 8.8% during competition) and a further 40.5% during endurance (30.2%) or strength training (10.2%); the remaining 22.4% occurred during other training, outside sport, or could not be clearly attributed (Table 3).

**Table 3:**
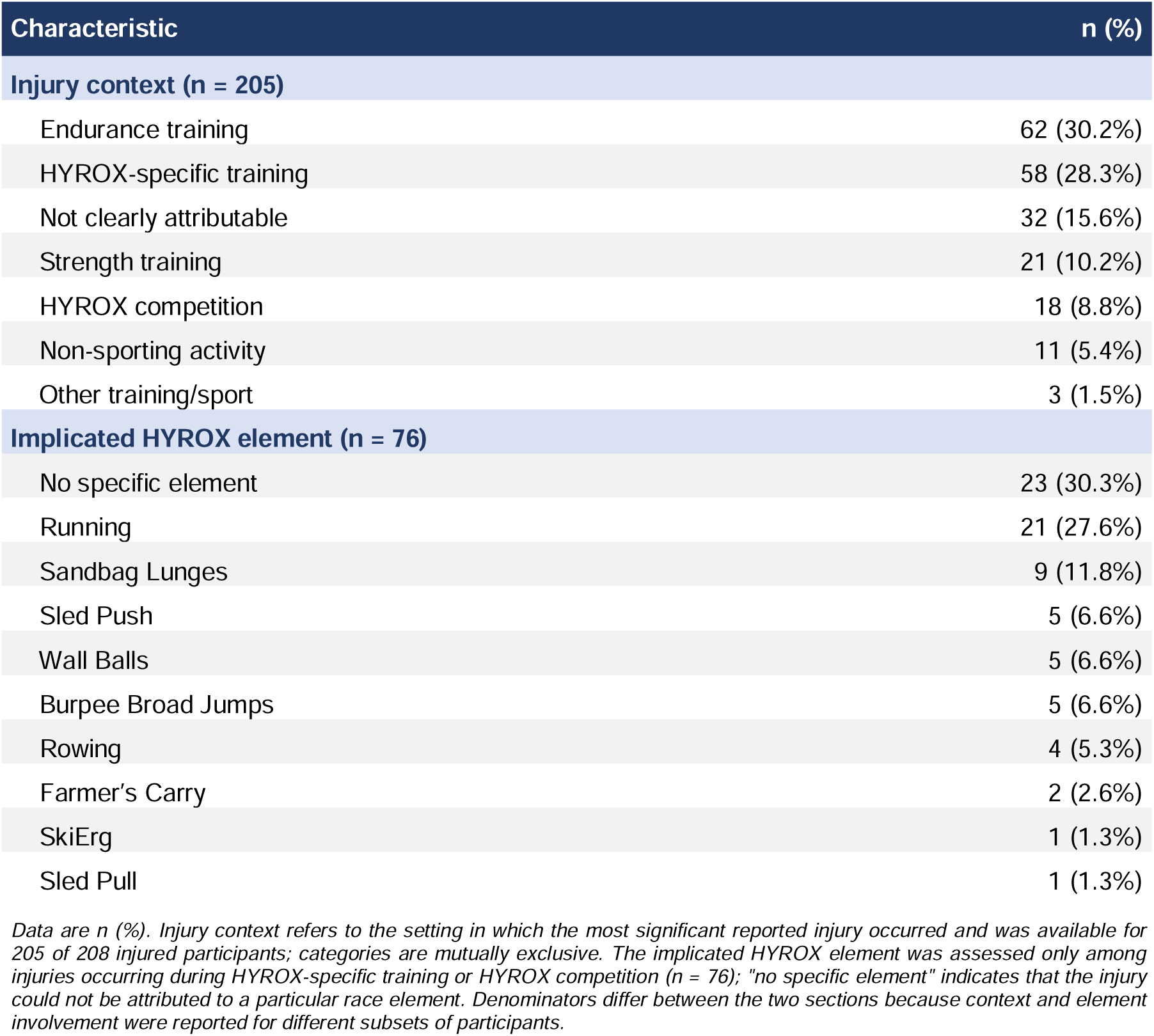
Injury context and implicated HYROX element.

Among the 76 injuries during HYROX-specific training or competition, 69.7% were attributed to a specific element – most often running (27.6%) and sandbag lunges (11.8%) – while no specific element was identified in 30.3% (Table 3).

In exploratory analyses restricted to injuries with complete context and injury- characterisation data, injuries attributed to endurance or HYROX-specific training were predominantly lower-extremity injuries (90/116; 78%) and had a gradual onset (75/116; 65%), whereas the small strength-training subgroup (n = 20) more often involved the spine/back (9/20; 45%) and had an acute onset (10/20; 50%).

### Factors associated with injury

In complete-case logistic regression (n = 413), higher HYROX-specific training frequency was the only factor independently associated with reporting at least one injury (adjusted odds ratio (OR) 1.61, 95% CI 1.20–2.16; p = 0.001). Crude associations for HYROX-specific training volume and self-rated intensity did not persist after adjustment and no other measured factor, including age, gender and BMI, was independently associated (Table 4). A complementary modified Poisson regression yielded a consistent association (prevalence ratio 1.24, 95% CI 1.10– 1.40).

**Table 4:**
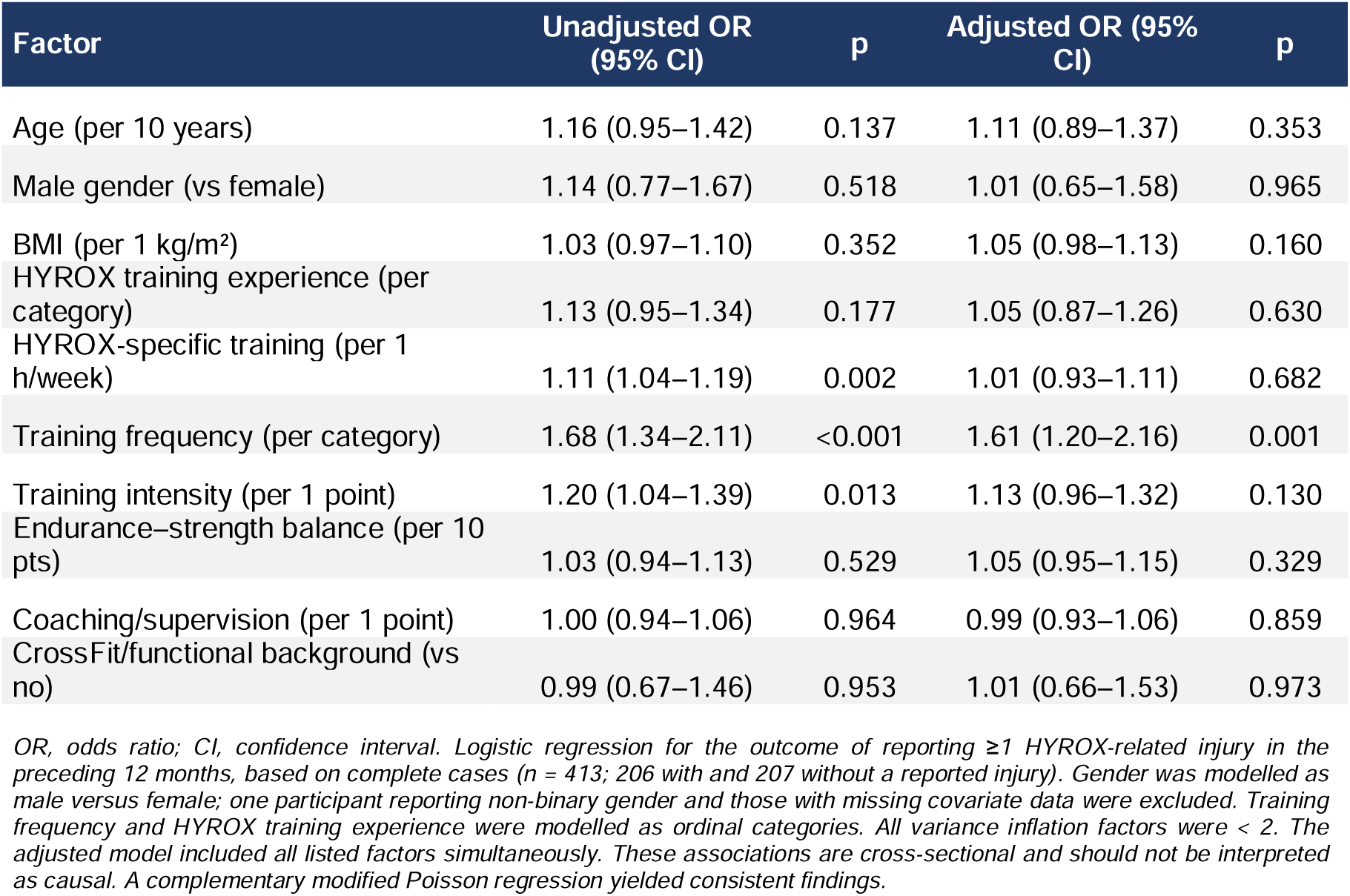
Factors associated with reporting ≥1 HYROX-related injury (n = 413)

The frequency association was consistent across prespecified sensitivity analyses: modelling HYROX-specific weekly volume as tertiles (OR 1.56, 95% CI 1.15–2.10), restricting the analysis to fully completed responses (n = 379; OR 1.49, 95% CI 1.08– 2.04), or restricting it to competition-experienced participants (n = 332; OR 1.56, 95% CI 1.13–2.14). A summary of our key findings is presented in Figure 2.

**Figure 2:**
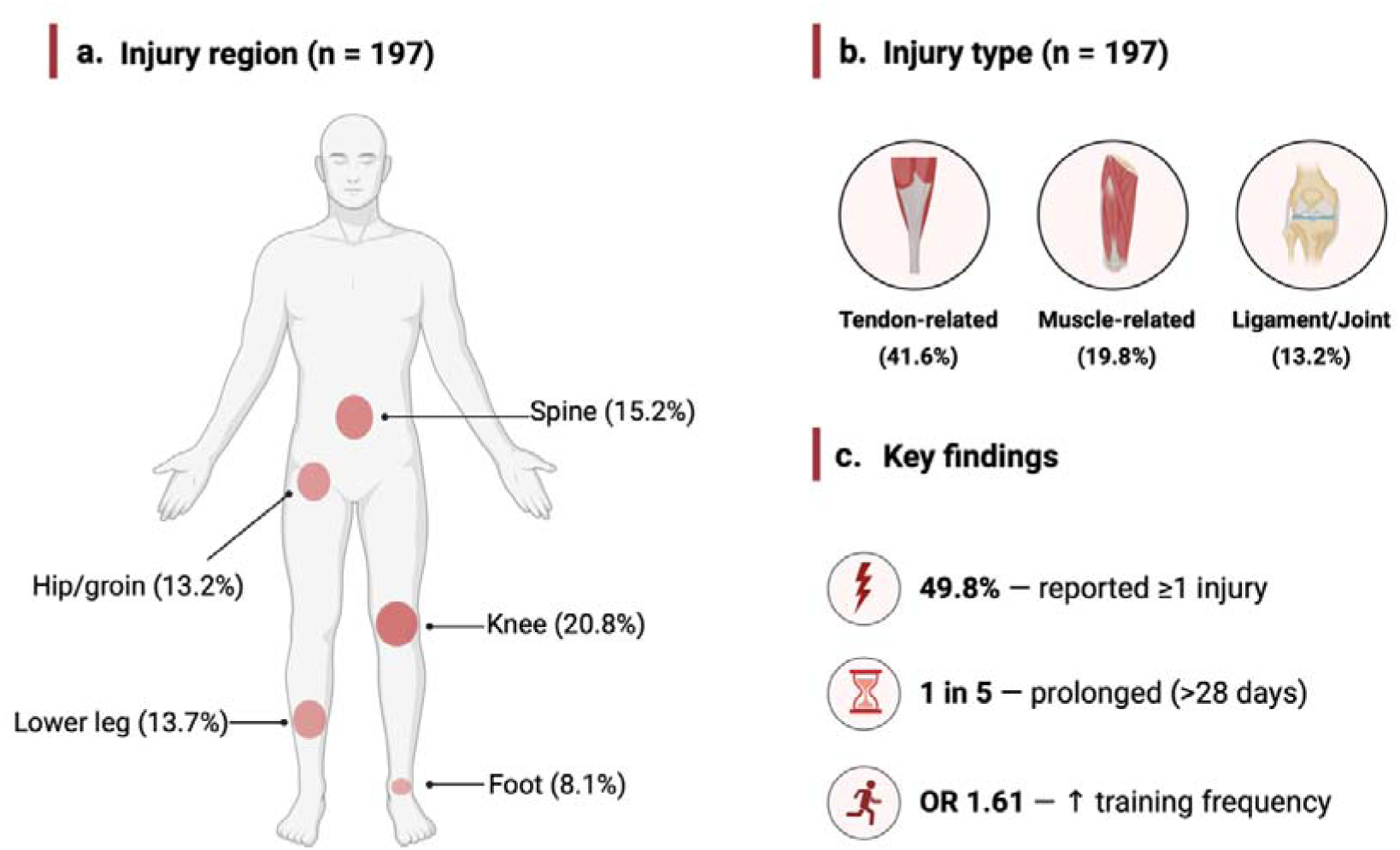
Central Figure. Summary of key injury findings in HYROX athletes. (a) Body region and (b) injury type of the most significant reported injury (n = 197); in (a) marker area is proportional to the frequency of injuries at each region. (c) Key findings: 12-month period prevalence of ≥1 HYROX-related injury 49.8% (208/418; 95% CI 45.0– 54.5); approximately one in five most significant injuries (20.3%) involved >28 days of training interruption or no return to the previous performance level; and higher HYROX-specific training frequency was the only measured factor independently associated with injury reporting (adjusted OR 1.61, 95% CI 1.20–2.16). Percentages are of the stated denominator. Findings are hypothesis-generating; the cross-sectional, self-reported design cannot establish cause or injury incidence.

## Discussion

This cross-sectional survey of 418 HYROX athletes provides, to our knowledge, the first and largest international epidemiological description of HYROX-related injuries to date. Over the preceding 12 months, almost half of participants (49.8%) reported at least one injury, with an exposure-adjusted lower-bound rate of 1.65 reported injuries per 1000 hours of total training exposure. Higher HYROX-specific training frequency was the only measured factor independently associated with injury reporting.

Because HYROX does not map neatly onto any single established sport, evidence from running, resistance training and related functional-fitness formats offers context but is not directly transferable.(3) A CrossFit systematic review reported a mean injury prevalence of 35.3% (range 12.8–73.5%).(4) Our estimate was higher than the mean but remained within the reported range. Given differences in injury definitions and study designs, these figures should be viewed as reference points rather than direct comparisons of injury risk.(4, 11) Similarly, the exposure-adjusted rate represents a lower-bound estimate rather than true incidence. Prospective surveillance with longitudinal exposure recording is needed to establish HYROX- specific injury incidence.

The observed injury profile was dominated by lower-extremity, tendon-related injuries of gradual or mixed onset. This profile closely resembles running-related injury patterns, in which the knee and lower leg are commonly affected and patellofemoral pain, Achilles tendinopathy and medial tibial stress syndrome are frequently reported.(14–17) Supporting this interpretation, running was the most frequently reported element among injuries attributed to a specific HYROX component. However, the prominence of spine/back injuries as the second most commonly affected region suggests that HYROX’s hybrid structure is also reflected in its injury distribution, as gym-based functional-fitness formats more commonly show shoulder- and spine-dominant patterns.(4, 18) Exploratory subgroup analyses showed a consistent pattern:(4, 18, 19) injuries attributed to endurance or HYROX-specific training were predominantly gradual-onset and lower-limb, whereas strength-training injuries more often involved the spine/back and had an acute onset. However, the small, non-prespecified subgroups make these findings hypothesis-generating.

Beyond anatomical distribution, the injuries had meaningful consequences for training: more than 90% of athletes with severity data reported at least one day of interrupted or reduced training, and approximately one in five reported >28 days of interruption or had not returned to their previous performance level. The broad, symptom-based definition therefore captured not only transient complaints but also injuries with prolonged effects on training and performance. About 70% of injured athletes sought medical or therapeutic care, indicating substantial healthcare utilisation. Future HYROX surveillance should quantify injury burden prospectively, including time-loss days per 1000 hours of exposure, in line with IOC consensus recommendations.(11)

Among the factors examined, only higher HYROX-specific training frequency showed an independent association with injury reporting; no other measured athlete- or training-related characteristic did. Findings from related sports on demographic risk factors are inconsistent, so the absence of independent associations with age, gender and BMI does not clearly diverge from the wider evidence base.(4, 18, 20, 21)

Crude associations for HYROX-specific training volume and self-rated training intensity did not persist after adjustment. Because training frequency, volume and self-rated intensity are closely interrelated, the present design cannot determine whether frequency itself, the distribution of training load across the week, or differences in recovery explain the observed association. Consensus work links loading frequency and insufficient recovery to overuse injury, including tendinopathy.(22, 23) Prospective HYROX studies should therefore examine how training frequency, load distribution and recovery jointly relate to injury over time.

### Limitations

This study has several limitations. First, the cross-sectional design cannot establish cause or temporal sequence; the frequency association could reflect reverse causation, as injuries may have altered subsequent training. Second, exposure and injuries were recalled over 12 months, risking under-reporting of minor complaints and less precise recall of onset and severity;(24) injuries were self-reported without clinical verification, and the broad, symptom-based definition limits comparability with studies using narrower time-loss or medical-attention definitions. For the 154 participants (36.8%) with less than 12 months of HYROX experience, the recall period exceeded their exposure window. Furthermore, the purpose-designed questionnaire was pilot-tested but not formally validated. Third, the open recruitment link precluded a response rate and could not fully exclude duplicate participation; injured or particularly engaged athletes may have been over-represented, and the sample was weighted towards experienced, competition-oriented participants and participants residing in Germany, limiting generalisability. Fourth, exposure was extrapolated from average weekly hours across 52 weeks and participants reporting more than one injury were conservatively counted as two events, so the exposure-adjusted rate is a lower-bound estimate; because HYROX-related injuries were counted against total (all-activity) rather than HYROX-specific exposure, it should not be interpreted as HYROX-specific incidence; relatedness to HYROX was self-attributed, and only 37.1% of injuries occurred during HYROX-specific training or competition. Fifth, detailed data covered at most two injuries per participant, mainly the most significant, so the injury profile may under-represent milder or recurrent complaints. Finally, although the 206 injured participants approached the number suggested for detecting small to moderate associations,(25) smaller associations with age, gender, BMI or experience may have remained undetected.

### Clinical and research implications

These findings are hypothesis-generating and cannot support causal or preventive recommendations, but they have practical relevance. The predominance of gradual-onset, lower-extremity and tendon-related complaints suggests that athletes, coaches and clinicians should be alert to overuse problems, particularly around running and lower-limb loading, and that overall training load – including weekly frequency – deserves attention. These findings highlight the need for prospective, exposure-based HYROX surveillance. Future studies should record activity-specific exposure and injury burden with standardised overuse-injury questionnaires,(26) and examine how training frequency, load distribution and recovery relate to injury risk and prevention.

### Conclusion

In this first international epidemiological study of HYROX athletes, HYROX-related injuries were common over 12 months (period prevalence 49.8%; exposure-adjusted lower-bound rate 1.65 reported injuries per 1000 hours of total training exposure). The most significant injuries were predominantly gradual-onset, lower-extremity and tendon-related, and a meaningful proportion led to prolonged training restriction. Higher HYROX-specific training frequency was the only measured factor independently associated with injury reporting. These findings provide an initial benchmark for HYROX injury epidemiology and support prospective studies to establish HYROX-specific incidence and burden. HYROX’s fixed, reproducible race structure makes it a useful real-world model for such research.

## Supplement

**Supplementary Table 1:** HYROX competition characteristics of the study sample (n = 418).

| Characteristic | n (%) |
| --- | --- |
| **Competition status (n = 418)** |  |
| Regular competitor | 180 (43.1%) |
| ≥1 competition completed | 156 (37.3%) |
| Planning first competition | 43 (10.3%) |
| Trains HYROX-style, never competed | 39 (9.3%) |
| **Competition formats (n = 336, multiple response)** |  |
| Doubles | 254 (75.6%) |
| Open (Individual) | 215 (64.0%) |
| Pro (Individual) | 103 (30.7%) |
| Relay | 59 (17.6%) |
| Elite | 17 (5.1%) |
| **Competitions completed (n = 336)** |  |
| 1 | 77 (22.9%) |
| 2 | 55 (16.4%) |
| 3–5 | 94 (28.0%) |
| 6–10 | 61 (18.2%) |
| > 10 | 49 (14.6%) |

Data are n (%). Competition status was assessed for all participants (n = 418); competition formats and number of competitions completed were assessed only among participants with competition experience (≥1 completed or regular competitor; n = 336). Competition formats were captured as a multiple-response item; categories are not mutually exclusive and percentages therefore sum to more than 100%.

**Supplementary Table 2:** HYROX background and training characteristics (n = 418).

| Characteristic | n = 418 |
| --- | --- |
| **HYROX background** |  |
| **HYROX training experience — n (%)** |  |
| < 6 months | 62 (14.8%) |
| 6–12 months | 92 (22.0%) |
| 1–2 years | 170 (40.7%) |
| 3–4 years | 56 (13.4%) |
| > 4 years | 38 (9.1%) |
| **Training characteristics** |  |
| Total training, h/week — median (IQR) | 8 (6–10) |
| HYROX-specific training, h/week — median (IQR) | 4 (2–6) |
| **Training frequency — n (%)** |  |
| < 1 day/week | 28 (6.7%) |
| 1–2 days/week | 158 (37.8%) |
| 3–4 days/week | 147 (35.2%) |
| 5–6 days/week | 74 (17.7%) |
| Daily | 11 (2.6%) |
| Training intensity (1–10) — median (IQR) (n = 417) | 7 (7–8) |
| Endurance–strength balance (0–100) — median (IQR) (n = 417) | 41 (28–60) |
| Coaching/supervision (1–10) — median (IQR) | 7 (2–9) |

Data are n (%) or median (IQR). IQR, interquartile range. Denominators are given where data were missing for individual variables. Total training volume refers to all training over the preceding 12 months; HYROX-specific training volume and frequency refer to HYROX-specific training only. Self-rated training intensity is scored from 1 (very low) to 10 (very high). Endurance–strength balance ranges from 0 (entirely endurance-focused) to 100 (entirely strength-focused). Coaching/supervision is scored from 1 (never) to 10 (always).

**Supplementary Table 3:**
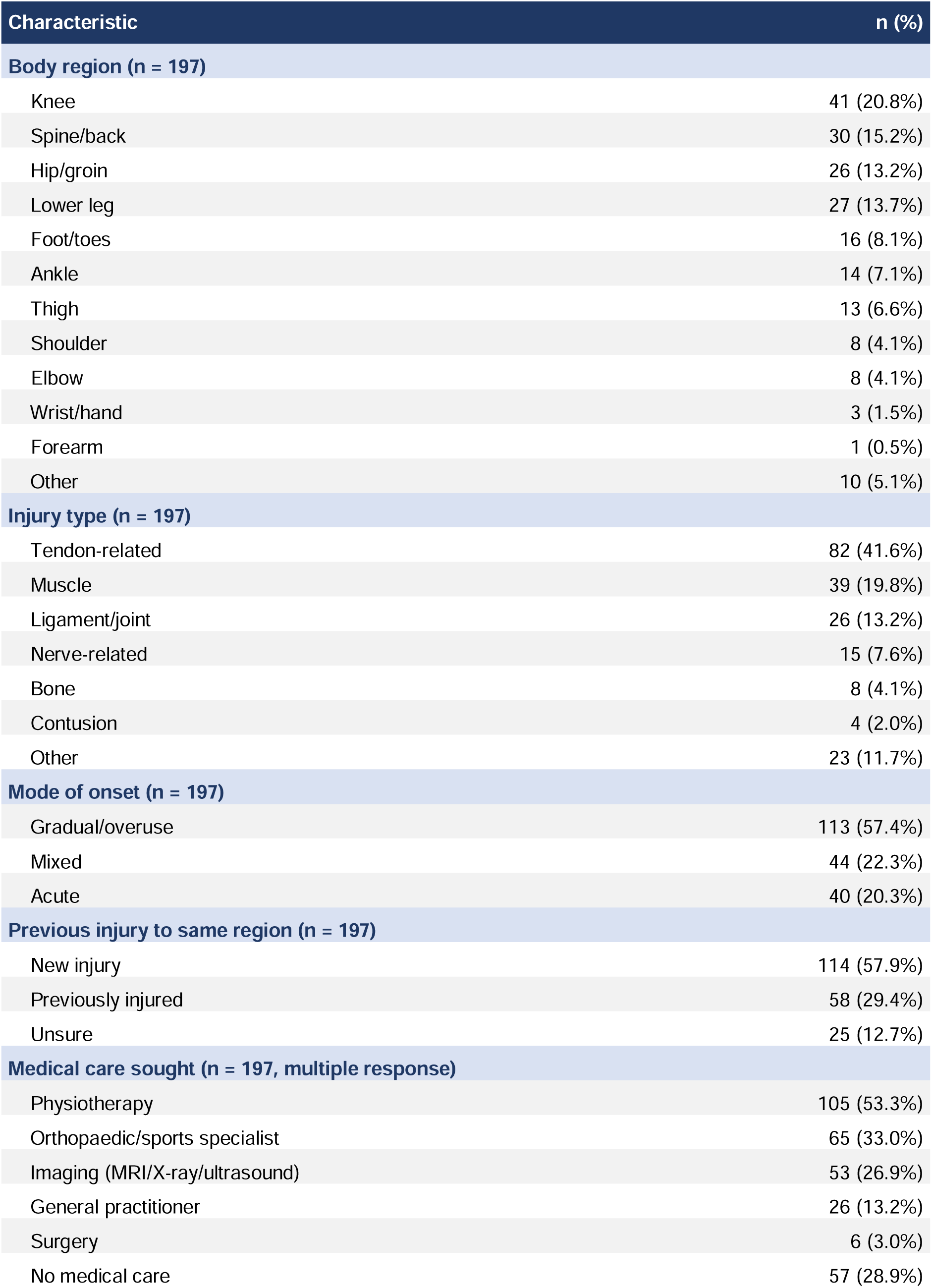
Characteristics of the most significant reported injury among injured participants (n = 197).

**Supplementary Table 4:** Characteristics of the second reported injury (n = 44)

| Characteristic | n (%) |
| --- | --- |
| **Body region** |  |
| Knee | 9 (20.5%) |
| Spine/back | 5 (11.4%) |
| Shoulder | 5 (11.4%) |
| Lower leg | 5 (11.4%) |
| Foot/toes | 5 (11.4%) |
| Hip/groin | 3 (6.8%) |
| Thigh | 3 (6.8%) |
| Wrist/hand | 2 (4.5%) |
| Ankle | 2 (4.5%) |
| Elbow | 1 (2.3%) |
| Other | 4 (9.1%) |
| **Injury type** |  |
| Tendon-related | 20 (45.5%) |
| Muscle | 8 (18.2%) |
| Ligament/joint | 6 (13.6%) |
| Bone | 2 (4.5%) |
| Nerve-related | 1 (2.3%) |
| Contusion | 1 (2.3%) |
| Other | 6 (13.6%) |
| **Mode of onset** |  |
| Gradual/overuse | 25 (56.8%) |
| Mixed | 9 (20.5%) |
| Acute | 10 (22.7%) |
| **Training interruption** |  |
| No interruption (0 days) | 6 (13.6%) |
| 1–3 days | 7 (15.9%) |
| 4–7 days | 10 (22.7%) |
| 8–28 days | 15 (34.1%) |
| > 28 days | 4 (9.1%) |
| No return to previous level yet | 2 (4.5%) |

Data are n (%). This table summarises the second injury reported by the 44 of 85 participants with more than one HYROX- related injury who provided second-injury details; 36 declined to provide further details and 5 had missing follow-up data. Body region, injury type and mode of onset were single-response items. Training interruption categories are mutually exclusive; "no return to previous level yet" denotes injuries from which participants had not returned to their pre-injury performance level by the time of survey completion. Because second-injury data were incomplete and did not capture the total number of injuries per participant, they were not pooled with the main injury-pattern analyses.

**Supplementary Figure 1:**
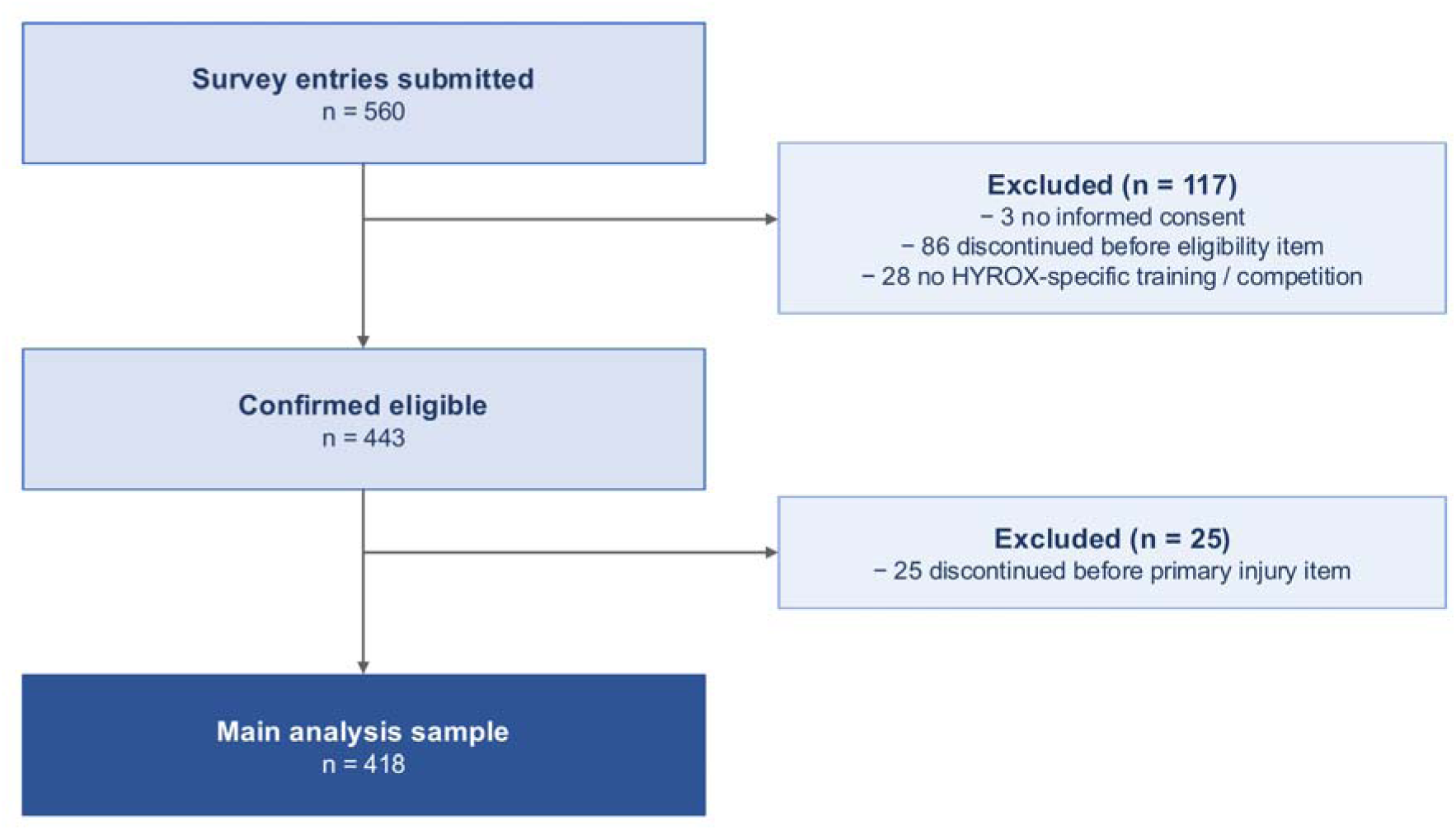
Flow of participants through the study, from survey submission to the main analysis sample.

## Declarations

### Authors’ contributions

C.K.: Conceptualization, Methodology, Investigation, Data curation, Formal analysis, Project administration, Writing – original draft.

L.K.: Investigation, Writing – original draft. M.B.: Methodology, Writing – review & editing.

P.Be.: Formal analysis, Supervision, Writing – review & editing. P.Bi.: Supervision, Writing – review & editing.

P.Z.: Supervision, Writing – review & editing. M.S.: Methodology, Writing – review & editing.

M.Z.: Conceptualization, Methodology, Writing – original draft, Writing – review & editing.

All authors reviewed and approved the final manuscript and accept responsibility for the integrity of the work. M.Z. is the guarantor.

## AI-Acknowledgement

The authors used Claude (Anthropic; version 1.18286.0, 2026) to assist with translation of the study questionnaire into English, language editing and review of statistical outputs; all authors reviewed and verified the content and take full responsibility for it.

## Ethics approval

The study was approved by the Ethics Committee of the Technical University of Munich (2026-32-S-CT; 3 February 2026) and conducted in accordance with the Declaration of Helsinki. Participation was voluntary and uncompensated; all participants provided electronic informed consent before completing the anonymous questionnaire. The participant information and consent text are provided in the online supplementary material.

## Consent for publication

Not applicable.

## Data availability statement

Anonymised participant-level data are not publicly available because participants consented to analysis and publication of results, not to public deposition of the dataset. Data may be available from the corresponding author on reasonable request, subject to institutional approval.

## Acknowledgements

None.

